# The Value of Admission Avoidance: Cost-Consequence Analysis of One-Year Activity in a Consolidated Service

**DOI:** 10.1101/2023.01.05.23284217

**Authors:** Carme Hernandez, Carme Herranz, Erik Baltaxe, Nuria Seijas, Rubèn González-Colom, Maria Asenjo, Emmanuel Coloma, Joaquim Fernandez, Emili Vela, Gerard Carot-Sans, Isaac Cano, Josep Roca, David Nicolas, JADECARE consortium

## Abstract

**Background:** Many advantages of hospital at home (HaH), as a modality of acute care, have been highlighted, but controversies exist regarding the cost-benefit trade-offs.

**Objective:** To assess health outcomes and analytical costs of hospital avoidance (HaH-HA) in a consolidated service with over ten years of delivery of HaH in Barcelona (Spain).

**Methods:** A retrospective cost-consequence analysis of all first episodes of HaH-HA, directly admitted from the emergency room (ER) in 2017–2018, was carried out. HaH-HA was compared with a propensity-score-matched group of contemporary patients admitted to conventional hospitalization (Controls). Mortality, re-admissions, ER visits, and direct healthcare costs were evaluated.

**Registration:** ClinicalTrials.gov (26/04/2017; NCT03130283).

**Results:** HaH-HA and Controls (n=441 each) were comparable in terms of age (73 [SD16] *vs* 74 [16]), gender (male, 57% *vs* 59%), multimorbidity, healthcare expenditure during the previous year, case mix index of the acute episode, and main diagnosis at discharge. HaH-HA presented lower mortality during the episode (0 vs. 19 (4.3%); *p* < 0.001). At 30 days post-discharge, HaH-HA and Controls showed similar re-admission rates; however, ER visits were lower in HaH-HA than in Controls (28 (6.3%) vs 34 (8.1%); p = 0.044). Average costs per patient during the episode were lower in the HaH-HA group (€ 1,078) than in Controls (€ 2,171). Likewise, healthcare costs within the 30 days post-discharge were also lower in HaH-Ha than in Controls (*p* < 0.001).

**Conclusions:** The study showed higher performance and cost reductions of HaH-HA in a real-world setting. The identification of sources of savings facilitates scaling of hospital avoidance.

**Funding:** This article was funded by JADECARE project- HP-JA-2019 - Grant Agreement n^<u>º</u>^ 951442 (2020-2023), a European Union’s Health Program 2014-2020.

**KEY POINTS:**

- Hospital at home, modality hospital avoidance, shows high potential for value generation in real-world settings by improving health outcomes and generating cost-savings.
- Positive outcomes of hospital avoidance seem associated to adequate management change and digital support.

## INTRODUCTION

Over the last twenty years, hospital at home (HaH) has reached maturity in various health systems worldwide, although some heterogeneities have been reported across sites [1–3]. This service is currently considered a consolidated alternative to inpatient care for selected patients requiring hospital admission [4]. Furthermore, HaH has shown high potential to promote continuity of care by preventing hospitalizations and reinforcing transitional care after discharge [5,6], thus enabling vertical integration between hospital and community-based care [7,8].

Despite the promising results and potential benefits associated with HaH, some controversies have been raised regarding the extent of the value generation in healthcare [9–11]. These discrepancies are partly explained by differences in the complexity of target patients and service delivery context, with important implications regarding the characterization of its different modalities, reimbursement regimes, and adoption strategies [12–14]. This heterogeneous scenario stresses the need for investigating real-world experiences in implementing and deploying HaH services.

In our center, a tertiary university hospital providing specialized care to a catchment population of 520,000 citizens, HaH was implemented in 2006 as a mainstream service across specialties covering two modalities of HaH: hospital avoidance (HaH-HA) and early discharge (HaH-ED) [7,15,16]. The service provides acute, home-based, short-term care aiming at either entirely replacing conventional hospitalization (hospital avoidance) or accelerating discharge (early discharge). This model was progressively implemented across the entire healthcare system in our region between 2011 and 2015, with preliminary positive results [7,17,18]. These positive results prompted the Catalan Health Service, the only public health payer providing universal healthcare to the 7.7 million population, to scale up the HaH service across the region and set a specific reimbursement model between 2016 and 2020 [19].

The long-lasting experience with HaH service and analytical accounting used in our center sets a privileged scenario to investigate the benefits and costs associated with this service. Therefore, we conducted a cost-consequence analysis (CCA) [14] of all first episodes of HaH-HA registered within a one-year course after more than one decade of implementation and consolidation of the HaH service in our center.

## METHODS

### Study groups and design

This was a retrospective CCA of all first-time HaH-HA admissions issued from the emergency room department in the Hospital Clínic of Barcelona among non-surgical patients between October 31, 2017, and November 1, 2018. The costs and outcomes of HaH-HA patients were compared with a 1:1 matched comparator group of conventional hospitalizations in our center. Patients under the modality HaH-ED were excluded from the analysis.

Candidates to HaH-HA were screened in the emergency room by trained professionals of the HaH team. Individuals were eligible for HaH-HA if: they were aged 18 years or older, lived in their house within the catchment area, had a formal or informal caretaker (including relatives) available 24 hours per day, had a phone at home and signed the informed consent to be hospitalized at home. We considered all medical conditions.

The comparator group (Controls) was built from non-surgical patients admitted for conventional hospitalization from the emergency room within the same period. We paired HaH-HA patients with control patients 1:1 using a propensity score matching (PSM) [20,21] and genetic-matching technique [22]. For matching purposes, we took into account two sets of matching variables to ensure patients’ comparability regarding both baseline characteristics (i.e., before admission) and hospitalization characteristics.

The first set of matching variables included age, gender, number of admissions in the previous year, patient’s healthcare costs across the health system in the previous year, and health risk based on the adjusted morbidity groups (AMG) index [23]. The AMG is a summary measure of morbidity that considers a weighted sum of all chronic and relevant acute conditions from all diagnostic group codes of the International Classification of Diseases, clinical modification (ICD-10-CM). The AMG can be used as a numerical index or as population-based risk groups, defined according to percentile thresholds for the distribution of the AMG index across the entire population of Catalonia. Both the index and the risk groups have shown a good correlation with relevant health outcomes and the use of healthcare resources [24,25].

The second set of variables for paring HaH-HA and control patients included relevant characteristics of the hospitalization episode, such as the main diagnosis at discharge based on the ICD-10-CM categories and the case mix index (CMI). The CMI summarizes the severity and complexity of the main diagnosis and health events occurring during the hospital stay.

### Characteristics of home and conventional hospitalizations

The HaH-HA group followed the standard of care for HaH at Hospital Clínic de Barcelona, which has been extensively reported elsewhere [7]. Briefly, a patient admitted to HaH-HA is assessed in person daily by the HaH team, which consists of either a nurse or a nurse and physician (at physician’s discretion) with remote access to the patient’s electronic record. Interventions available at home include regular tests (e.g., blood and microbiology tests, clinical ultrasound, electrocardiogram), most of the intravenous and nebulized treatments, and oxygen therapy. A pathway for elective transfer back to the hospital (e.g., for additional tests not available at home) and emergency transfer in case of clinical deterioration are also available.

The control group followed the usual care for in-house hospitalizations; patients were assigned to a hospital bed within the corresponding service according to the primary diagnosis and followed up by the medical and nurse staff of the corresponding ward or service.

Upon discharge, patients in the two groups were transferred to the corresponding primary care teams, with access to electronic health records. However, the HaH team shares responsibilities with the primary care team during the transitional care period until 30 days after discharge.

### Outcomes and costs

The CCA included health outcomes and direct costs [26]. Health outcomes included length of hospital stay, 30-day mortality, and all-cause hospital admissions and visits to the ER within the 30 days following discharge. In patients admitted to HaH-HA, we also collected the patient experience by administering a 9-item satisfaction questionnaire [7] on discharge.

Costs were estimated using an analytical accounting approach [27]. Direct costs included honoraria of staff professionals, pharmacological and non-pharmacological therapy, consumables, testing and procedures, transportation, catering, and structural costs (i.e., information systems, electricity, etc). We also considered healthcare expenditure associated with any resource use of the healthcare system during the 30 days following discharge.

The two data sources used for the study were: the SAP Health Information System at HCB and the Catalan Health Surveillance System (CHSS) for analysis of the acute episode and calculations after discharge, respectively.

### Data analysis

Health outcomes and costs were described by the number and percentage over available data for categorical variables and mean and standard deviation (SD), or median and interquartile range (IQR, defined by the 25^th^ and 75^th^ percentiles), as appropriate. The matching parameters were tuned to enhance the covariate balancing, as follows: caliper: 0.2, function: logit, replace: FALSE, ratio: 1:1, matching method: Genetic Matching. Genetic Matching uses an optimization algorithm based on “GENetic Optimization Using Derivatives (GENOUD)” [28] to check and improve covariate balance iteratively, and it is a generalization of propensity score and Mahalanobis distance [29]. The matching was assessed by the Mahalanobis distance, Rubin’s B (the absolute standardized difference of the means of the linear index of the propensity score in the HaH-HA and Controls) and Rubin’s R [30] (the ratio of HaH-HA to Controls variances of the propensity score index) metrics. Quality of comparability between HaH-HA and Controls after PSM was considered acceptable if Rubin’s B was less than 0.25 and Rubin’s R was between 0.5 and 2. Unpaired Student T tests, Mann-Whitney, and Chi-squared tests comparing HaH-HA with Controls were used to assess changes in the costs and clinical outcomes. Data analyses were conducted using R [31], version 3.6.1 (R Foundation for Statistical Computing, Vienna, Austria). The threshold for significance was set at a two-sided alpha value of 0.05.

### Ethical conduct of the study

The Ethical Committee for Human Research at Hospital approved the study protocol (refs. 2017-0451 and 2017-0452), which waived the collection of informed consent for the secondary use of routine care data. All methods were conducted in accordance with the relevant guidelines and regulations, including the General Data Protection Regulation 2016/679 on data protection and privacy for all individuals within the European Union and the local regulatory framework regarding data protection.

## RESULTS

### Study participants

During the study period, the ER department dictated 586 unplanned non-surgical HaH-HA admissions in patients without previous episodes of HaH. The comparator group was built using a dataset of 2,631 conventional non-surgical admissions carried out during the study period. After propensity score matching, the two groups: HaH-HA and Controls, consisted of 441 cases each (**Figure 1**).

**Figure 1.**
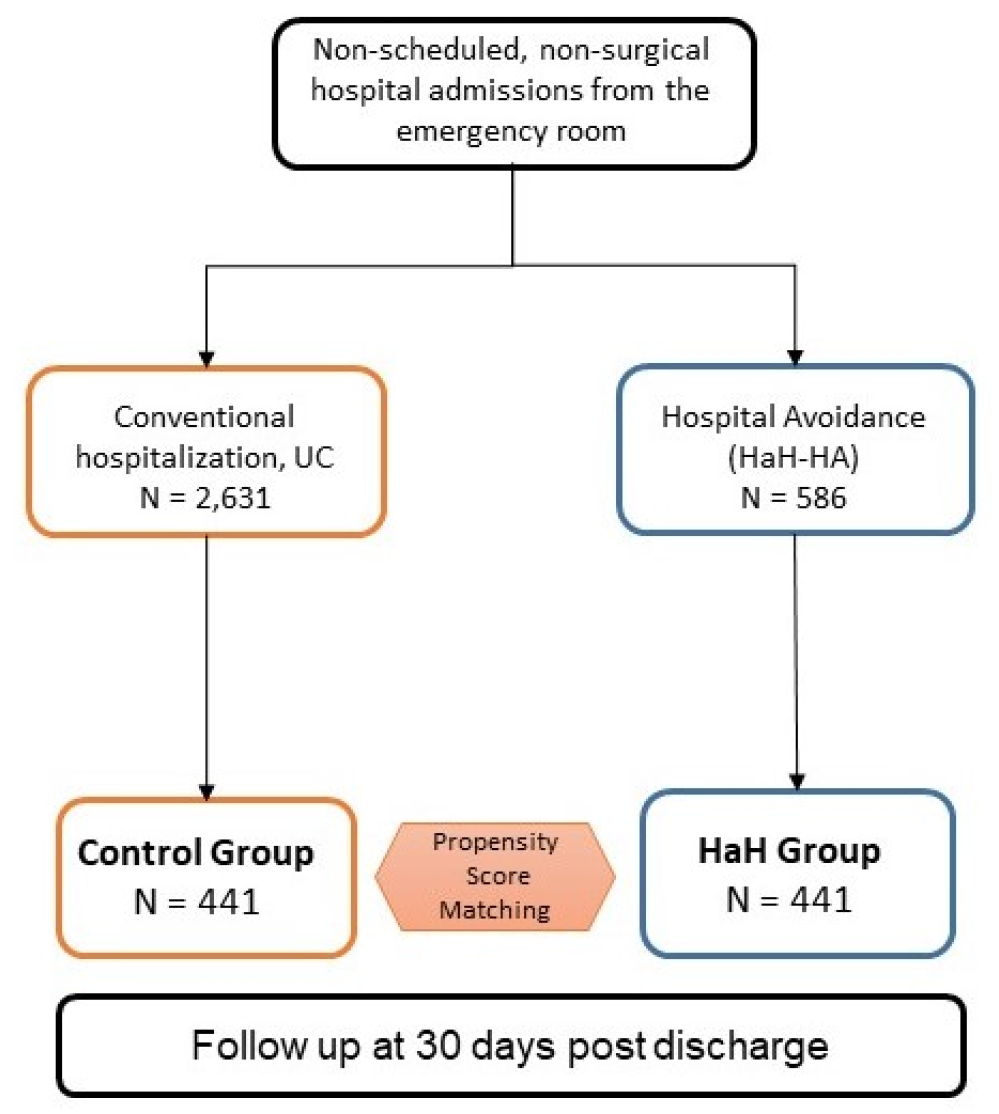
Number and distribution of patients. **Legend** – Five-hundred eighty-six first episodes of HaH admissions, directly from the Emergency Room (HaH-HA), were registered during the study period. After propensity score matching, the HaH-HA group fell to 441 patients (Comparisons among the two study groups and the study population of 586 patients are reported in **Tables 1S, 3S** and **4S**, see text for details).

We found no significant differences between the characteristics of the HaH-HA selected for the propensity-score matching (n=441) and those of the entire series of patients admitted to HaH-HA within the investigated period (n=586) (**Table S1**).

**Table 1** summarizes the baseline characteristics (i.e., before admission) of individuals included in the HaH-HA and the comparator group. The two groups were well balanced regarding their demographic characteristics and previous use of hospital resources and healthcare expenditure. The health risk on admission, measured using the AMG index, was also similar between groups. However, the stratification of patients across the population-based categories of health risk showed that the HaH-HA had a higher percentage of individuals in the intermediate-risk group and a lower percentage of individuals in the high-risk group than the control group.

**Table 1.** Characteristics of the study groups after propensity score matching before admission. **Legend.** PSM, propensity score matching; HaH-HA, Hospital al Home-Hospital Avoidance; Controls, Conventional hospitalizations; GMA, Adjusted Morbidity Groups scoring; *Matching variables.

|  | HaH-HA | Controls | P value |
| --- | --- | --- | --- |
| **SOCIO-DEMOGRAPHICS** |  |  |  |
| Age (years), mean (SD)* | 72.71 (16.30) | 73.94 (16.01) | 0.259 |
| Gender (male), n (%) | 250 (56.69) | 262 (59.41) | 0.412 |
| **USE OF HEALTH CARE RESOURCES** |  |  |  |
| **Hospital resources in previous 12 months** |  |  |  |
| Rate of all-cause emergency room visit, mean (SD) | 1.63 (1.04) | 1.75 (1.26) | 0.829 |
| Rate of all-cause Hospital admissions, mean (SD)* | 1.66 (1.22) | 1.62 (1.30) | 0.786 |
| Rate of planned admissions, mean (SD) | 1.37 (0.72) | 1.40 (0.87) | 0.832 |
| Last visit (days) to outpatient clinic before admission, mean (SD) | 85.98 (91.96) | 91.39 (94.39) | 0.522 |
| Last hospitalisation (days) before admission, mean (SD) | 192.16 (108.75) | 175.22 (126.70) | 0.262 |
| Length of stay in days (total days per year), mean (total) | 11.48 (1,538) | 11.49 (1,333) | 0.786 |
| Intensive care unit stays, n (%) | 19 (8.50) | 18 (9.60) | 0.547 |
| Outpatient visits, mean (SD) | 5.99 (7.19) | 5.45 (5.69) | 0.357 |
| **Hospital resources in previous 7 days** |  |  |  |
| Outpatient visits, mean (SD) | 1.11 (0.42) | 1.14 (0.42) | 0.765 |
| **Healthcare costs across tiers in previous year** |  |  |  |
| € per year, mean (SD)* | 5,627 (8,119) | 6,543 (6,869) | 0.070 |
| **MULTIMORBIDITY & SEVERITY** |  |  |  |
| GMA scoring, mean (SD)* | 24.94 (15.17) | 25.09 (14.51) | 0.884 |
| GMA category, n (%) |  |  |  |
| Tier 1 < P<sub>50</sub> | 6 (1.4) | 8 (1.8) | 0.798 |
| Tier 2 [P<sub>50</sub> - P<sub>80</sub>) | 31 (7.0) | 30 (6.8) | 0.988 |
| Tier 3 [P<sub>80</sub> - P<sub>95</sub>) | 97 (22.0) | 69 (15.7) | **0.019** |
| Tier 4 [P<sub>95</sub> - P<sub>99</sub>) | 83 (18.8) | 117 (26.5) | **0.005** |
| Tier 5 ≥ P<sub>99</sub> | 224 (50.8) | 217 (49.2) | 0.689 |
| **Statistics to assess matching** |  |  |  |
| Rubin's B |  |  | 0.2 |
| Rubin's R |  |  | 1.2 |

### Hospitalization characteristics and outcomes

The main diagnosis at discharge in the two groups, HaH-HA and Controls, showed the same distribution of percentages across the ICD-10-CM disease groups: 25% were urinary tract infections, 15% chronic respiratory diseases, 13% pneumonia, 11% acute lower respiratory tract infections, 9% heart failure, 8% skin infections, 6% flu, 3% symptoms, injury, and poisoning, 3% hypertensive disease and other heart diseases, 3% pneumonitis caused by bronchial aspiration, 4% other conditions requiring admission. Detailed information is provided in **Table 2S**.

**Table 2.** Characteristics of the acute episode and main outcomes. **Legend.** HaH-HA, Hospital at Home-Hospital Avoidance; Controls, Conventional hospitalizations; N/A, not applicable

|  | HaH-HA<br>(n=441) | Controls<br>(n=441) | P value |
| --- | --- | --- | --- |
| **Total length of stay (days), mean (SD)** | 7.89 (4.37) | 7.37 (6.17) | 0.142 |
| **Case Mix Index** | 0.69 | 0.73 | 0.633 |
| **Use of resources during Hospital Avoidance** |  |  |  |
| All-cause Emergency Room visits, n (%) | 6 (1.36) | N/A | N/A |
| All-cause In-Hospital re-admissions, n (%) | 18 (4.08) | N/A | N/A |
| **Mortality during episode, n (%)** | 0 (0) | 19 (4.31) | **N/A** |
| **Outcomes at 30 days after discharge** |  |  |  |
| All-cause Emergency Room visits, n (%) | 28 (6.35) | 34 (8.06) | **0.044** |
| All-cause Hospital admissions |  |  |  |
| Unplanned Hospital admissions, n (%) | 24 (5.44) | 23 (5.45) | 0.777 |
| Planned admissions, n (%) | 13 (2.95) | 10 (2.37) | 0.598 |
| Mortality, n (%) | 7 (1.59) | 7 (1.66) | 0.933 |

The characteristics of the acute hospitalization episode are summarized in **Table 2**. The two groups had similar CMI and length of stay. However, mortality and visits to the emergency room for any cause within the 30 days following discharge were significantly higher among patients with conventional hospitalization). In the HaH-HA group, 6 (1.4%) patients worsened their clinical condition during the episode, requiring a visit to the ER department and returning home. Likewise, 18 (4.1%) patients discontinued HaH-HA for similar reasons and were admitted to conventional hospitalization. The administration of the satisfaction questionnaire to patients admitted to HaH-HA revealed that 97% were highly satisfied with the service (**Figure S1**). Comprehensive information on the acute episode is provided in **Table 3S**.

Mortality and hospital admissions for any cause within the 30 days following discharge were similar in the two groups (**Table 2**). However, the conventional hospitalization group reported a significantly higher percentage of all-cause visits to the emergency room within the 30-day post-discharge period.

### Healthcare costs

The total direct costs associated with the hospitalization episodes were € 475k and € 957k for the HaH-HA and comparator groups, respectively. **Figure 2** displays the cost per patient (average according to concepts and cost distribution across each group). In the two groups, costs associated with staff salaries accounted for the greatest proportion of all items. The average cost per episode was € 1,078 and € 2,171 (*p* < 0.001) for HaH-HA and conventional hospitalization episodes, respectively. Cost savings per episode in HaH-HA compared to conventional hospitalization were mostly attributable to staff (€ 867 *vs.* € 1,539; *p* < 0.001), followed by catering (€ 0 *vs.* € 149), infrastructure (€ 13 *vs.* € 151; P<.001), testing (€ 21 *vs.* € 124; *p* < 0.001), and consumables (€ 31 *vs.* € 89; *p* < 0.001). HaH-HA had no statistically significant impact on costs associated with the treatment (€ 110 *vs* € 119; *p* = 0.662). Contrarily, compared to usual care, HaH-HA showed significantly increased costs on staff transportation (€ 36 *vs.* € 0). In the HaH-HA group, none of the patients or their relatives required additional external support during the hospitalization episode. Transportation to the hospital, when needed, was afforded by the public healthcare payer.

**Figure 2.**
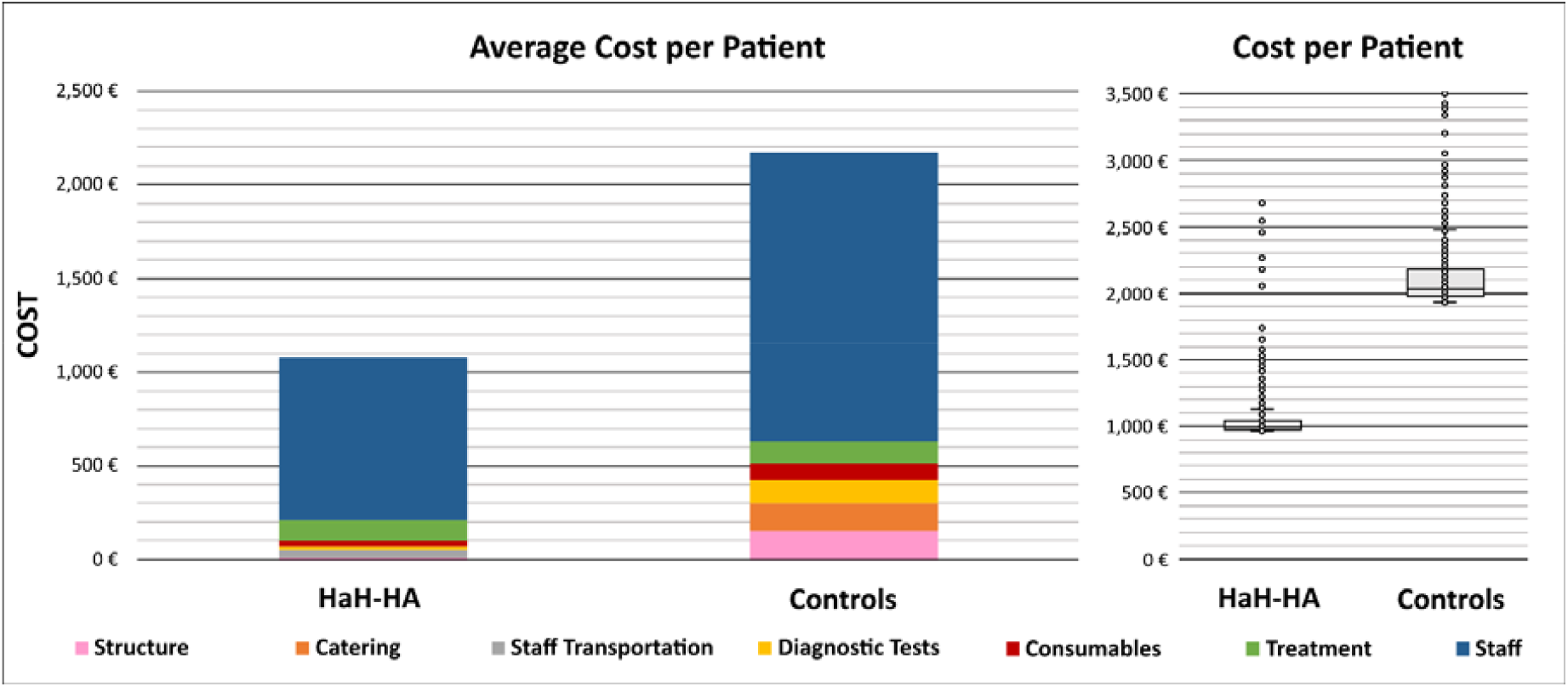
Average cost per pacient. **Legend** – In the left panel, the two columns indicate the average cost per patient for HaH-HA (Hospital at Home-Hospital Avoidance) and matched Controls (conventional hospitalizations), respectively. The colors indicate the weight of the different cost components (see text for details). The right panel depicts the box plots of the cost analysis.

The general healthcare expenditure within the 30 days following discharge was significantly lower in individuals admitted to HaH-HA than those with conventional hospitalization (€ 764 vs. € 1,022; P<0.001). For the two groups, all post-discharge healthcare costs were afforded by the public healthcare payer; no indirect costs afforded by patients or their relatives were considered in the analysis. **Table S4** provides a detailed list of costs associated with healthcare resource consumption within the 30 days following discharge.

## DISCUSSION

In this control-matched comparison of HaH-HA and conventional hospitalization, we found that HaH-HA was associated with significantly lower mortality during hospitalization and lower visits to the emergency room within the 30 days following discharge, despite the similar characteristics of the two groups, including the patients’ CMI on admission and the length of stay. The overall cost per episode was nearly half in the HaH-HA compared with conventional hospitalization. This cost reduction was primarily attributed to staff, catering, infrastructure, and testing. Likewise, patients admitted for a HaH-HA showed significantly lower healthcare expenditure within the 30 days following discharge.

Our findings are in the upper range of care quality of HaH studies in Europe [32,33] US [34–36], and Australia [37–40], taking into account the heterogeneity of services in the type of care, reimbursement regimes, and adoption strategies. Furthermore, the maturity of both integrated care and digital support may strongly influence the success of implementation and adoption strategies [16]. This factor, which increases the complexity of assessing HaH, may have also influenced the cost assessment. Thus, the innovation and change management with digital support of the service, which was gradually implemented in the early phases of HaH [7,41] but accelerated during the study period, may have contributed to cost reduction and improved health outcomes observed in our analysis.

Using a control-matched population strengthens our analysis admitted for conventional hospitalization within the same period in a real-world setting. This approach required ruling out 145 patients of the 586 hospitalized at home within the investigated period. However, our analysis of the baseline characteristics showed no differences with the final analysis dataset; therefore, we do not expect this exclusion to limit the representativeness of our cohort. Other strengths of our analysis include the possibility of collecting integrated data regarding healthcare resource utilization (including primary care) before and after the hospitalization episode, as well as using of analytical accounting for the cost analysis. This approach provided a detailed picture of costs, which is impossible with case-mix payment tools, such as the diagnostic risk groups used in several reports.

On the other hand, the current study was limited to the assessment of the direct costs of the healthcare provider, losing sight of indirect costs (e.g., home careers, etc…). More importantly, we could not gather societal costs or economic burdens for caretakers or patients’ relatives. Therefore, our cost-consequence analysis from the healthcare provider and healthcare system perspective shall be expanded in the future by including all these indirect and societal costs.

Aside from highlighting the need for a more comprehensive analysis of costs, our study paves the way to identifying key performance indicators that consider both site-specific and general features and allow for continuous monitoring of HaH performance. Another aspect of HaH to be explored is the implications of this type of care for improving the continuity of care by fostering vertical integration (i.e., between specialized and community-based care) and horizontal integration (i.e., between healthcare and social care). Although the role of HaH in these integrations was out of the scope of our analysis, health professionals working in a HaH are a natural bridge between specialized and community-based care during the transitional period during and after discharge [42]. Hence, HaH should be promoted as a facilitator of integrated care pathways, and future studies should investigate the contribution of HaH to maintaining the continuity of care in these transitions.

## CONCLUSIONS

Our analysis indicates that HaH-HA adds overall value for healthcare providers and the healthcare system. Taken together, our study and the current literature on the field highlight the need for adopting comprehensive and applicable assessment frameworks for HaH to facilitate the comparability and transferability of the service.

## Data Availability

The datasets generated and/or analyzed during the current study are not publicly available due to administrative reasons but are available from the corresponding author upon reasonable request.

## FUNDING

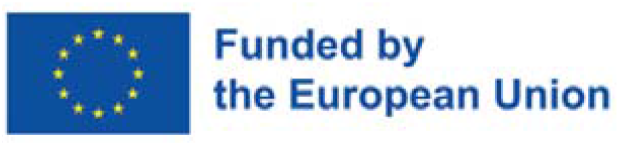

This article was funded by JADECARE project-HP-JA-2019 – Grant Agreement nº 951442 a European Union’s Health Program 2014-2020.

## Author Contributions

RG, IC, MCH, EV, MA, EB, NS, EC, JF, DN, JR and CH contributed to the preparation of this manuscript. RG, IC, MCH, EV, MA, EB, NS, EC, JF, DN, GC, JR and CH reviewed the full assessment report as well as this article, and can act as a guarantor for the overall content. MCH, GC, JC, RG and CH contributed to the sections relating to development of guidance and consultation and reviewed the manuscript for accuracy.

