## Supplementary material for "The Value of Admission Avoidance: Cost-Consequence Analysis of One-Year Activity in a Consolidated Service": Online supplement

*Hernandez C. et al (on-line supplementary material)*

**EXPANDED RESULTS – CHARACTERISTICS OF THE STUDY GROUPS**

**Table 1S** displays baseline data of the entire HaH-HA population (n=586), the HaH-HA group (n= 441) selected after Propensity Score Matching (PSM) and the corresponding Controls, conventional hospitalizations.

**Table 2S** informs on the distribution of main diagnostics, ICD-10 coding, in HaH-HA and Controls.

**Table 3S** provides data during the acute episode and during the 30-day period after discharge for the same datasets described in **Table 1S**.

**Table 4S** depicts information on operational costs of the service during the acute episode, as well as the community-based expenditure during the 30-day period after discharge following the same organization of the information indicated for **Tables 1S** and **3S**.

**Figure 1S** depicts the mean scoring for the different items of the questionnaire on patients/careers satisfaction administered to the HaH-HA patients at discharge.

**TABLE 1S.** Characteristics of the study population (n=586) and study groups (n=441 each) after propensity score matching, before admission. Only the p-values ≤ 0.05 have been displayed.

|  | **Study Population (n=586)** | **HaH-HA (n=441)** | **p value** | **Controls (n=441)** | **p value** |
| --- | --- | --- | --- | --- | --- |
| **SOCIO-DEMOGRAPHICS** |  |  |  |  |  |
| Age (years), mean (SD)* | 72.12 (16.32) | 72.71 (16.3) |  | 73.94 (16.01) |  |
| Gender (male), n (%)* | 334 (57.00) | 250 (56.69) |  | 262 (59.41) |  |
| **USE OF HEALTH CARE RESOURCES** | | | | | |
| **Hospital resources in previous 12 months** |  | | |  | |
| Rate of all-cause emergency room visit, mean (SD) | 1.71 (1.14) | 1.63 (1.04) |  | 1.75 (1.26) |  |
| Rate of all-cause Hospital admissions, mean (SD)* | 1.69 (1.13) | 1.66 (1.22) |  | 1.62 (1.3) |  |
| Rate of planned admissions, mean (SD) | 1.40 (0.70) | 1.37 (0.72) |  | 1.40 (0.87) |  |
| Last visit (days) to outpatient clinic before admission, mean (SD) | 79.34 (88.24) | 85.98 (91.96) |  | 91.39 (94.39) |  |
| Last hospitalisation (days) before admission, mean (SD) | 190.26 (108.14) | 192.16 (108.75) |  | 175.22 (126.70) |  |
| Length of stay in days (total days per year), mean (total) | 11.51 (2129) | 11.48 (1538) |  | 11.49 (1333) |  |
| Intensive care unit stays, n (%) | 27 (8.7) | 19 (8.5) |  | 18 (9.6) |  |
| Outpatient visits, mean (SD) | 6.41 (7.46) | 5.99 (7.19) |  | 5.45 (5.69) |  |
| **Hospital resources in previous 7 days** |  | | |  | |
| Outpatient visits, mean (SD) | 1.2 (0.55) | 1.11 (0.42) |  | 1.14 (0.42) |  |
| **Healthcare costs across tiers in previous year** |  | | |  | |
| € per year, mean (SD)* | 6,664.41 (8,738.58) | 5,627.06 (8118.86) |  | 6,543.05 (6868.5) |  |
| **MULTIMORBIDITY & SEVERITY** | | | | | |
| GMA scoring , mean (SD)* | 26.55 (15.82) | 24.95 (15.17) |  | 25.09 (14.51) |  |
| GMA category, n (%) |  | | |  | |
| *Tier 1 < P_50_* | 16 (3.63) | 6 (1.36) |  | 8 (1.81) |  |
| *Tier 2 [P_50_ - P_80_)* | 47 (10.66) | 31 (7.03) |  | 30 (6.8) |  |
| *Tier 3 [P_80_-P_95_)* | 83 (18.82) | 97 (22.00) |  | 69 (15.65) |  |
| *Tier 4 [P_95_-P_99_)* | 119 (26.98) | 83 (18.82) |  | 117 (26.53) |  |
| *Tier 5 ≥ P_99_* | 321 (72.79) | 224 (50.79) |  | 217 (49.21) |  |

**Legend.** CCA, Cost Consequence Analysis; Study population: all patients included in Hospital at Home-Hospital Avoidance (HaH-HA); HaH-HA, corresponds to the study population after propensity score matching (PSM); Controls, conventional hospitalizations, after PSM; GMA, Adjusted Morbidity Groups scoring; * Matching variables.

**Table 2S.** Distribution of main diagnosis at discharge.

| **Disease Category** | | **Study Group** | |
| --- | --- | --- | --- |
| **ICD 10** | **ICD Description** | **HH** | **Controls** |
| ***Urinary tract infection*** | | **117** | **117** |
| **N10** | *Acute tubulo-interstitial nephritis* | 26 | 15 |
| **N17** | *Acute kidney failure* | 1 | 12 |
| **N39** | *Other disorders of urethra and urinary tract* | 70 | 70 |
| **N41** | *Inflammatory diseases of prostate* | 17 | 8 |
| **N45** | *Orchitis and epididymitis* | 3 | 12 |
| ***Chronic lower respiratory diseases*** | | **67** | **67** |
| **J42** | *Chronic bronchitis* | 12 | 15 |
| **J43** | *Emphysema* | 7 | 9 |
| **J45** | *Asthma* | 7 | 7 |
| **J47** | *Bronchiectasis* | 9 | 14 |
| **J44** | *Other chronic obstructive pulmonary disease* | 32 | 22 |
| ***Pneumonia*** | | **55** | **55** |
| **J12** | *Viral pneumonia* | 0 | 2 |
| **J13** | *Pneumococcal pneumonia* | 4 | 9 |
| **J15** | *Bacterial pneumonia, not elsewhere classified* | 1 | 7 |
| **J18** | *Pneumonia, unspecified organism* | 50 | 37 |
| ***Acute lower respiratory infection and other respiratory disorders*** | | **46** | **46** |
| **J96** | *Other diseases of lung* | 3 | 6 |
| **J98** | *Other diseases of respiratory system* | 8 | 16 |
| **J20** | *Acute bronchitis* | 2 | 3 |
| **J22** | *Unspecified acute lower respiratory infection* | 32 | 18 |
| **J84** | *Other interstitial pulmonary diseases* | 1 | 3 |
| ***Hearth failure (I50)*** | | **37** | **37** |
| ***Infections of the skin and subcutaneous tissue*** | | **33** | **33** |
| **L02** | *Cutaneous abscess, furuncle and carbuncle* | 1 | 1 |
| **L03** | *Cellulitis and acute lymphangitis* | 29 | 29 |
| **S81** | *Open wound of knee and lower leg* | 1 | 0 |
| **S70** | *Superficial injury of hip and thigh* | 0 | 1 |
| **L97** | *Chronic ulcer of skin* | 2 | 2 |
| ***Flu (J10,J09,J11)*** | | **25** | **25** |
| ***Symptoms, injury and poisoning*** | | **16** | **16** |
| **R68** | *General symptoms* | 3 | 1 |
| **R78** | *Find of drugs and other substnces, not normally found in blood* | 10 | 5 |
| **T82** | *Complications of cardiac and vascular prosth dev/grft* | 0 | 3 |
| **T81** | *Complications of procedures, not elsewhere classified* | 0 | 4 |
| **R50** | *Fever of unknown origin* | 1 | 3 |
| **T83** | *Complications of genitourinary prosth dev/grft* | 1 | 0 |
| **T85** | *Complications of internal prosth dev/grft* | 1 | 0 |
| ***Hipetensive diseases and other heart diseases*** | | **15** | **15** |
| **I11** | *Hypertensive heart disease* | 1 | 0 |
| **I13** | *Hypertensive heart and chronic kidney disease* | 4 | 0 |
| **I21** | *Acute myocardial infarction* | 1 | 5 |
| **I20** | *Angina pectoris* | 0 | 1 |
| **I26** | *Acute pulmonary heart disease* | 3 | 4 |
| **I48** | *Cardiac dysrhythmias* | 1 | 4 |
| **I82** | *Other venous embolism and thrombosis* | 5 | 1 |
| ***Pneumonitis caused by bronchial aspiration (J69)*** | | **13** | **13** |
| ***Neutropenia and anemia (D61, D70)*** | | **5** | **5** |
| ***Infections (A41, A09, B97)*** | | **2** | **2** |
| ***Malignant neoplasms (C34, C67)*** | | **2** | **2** |
| ***Other*** | | **8** | **8** |
| **K05** | *Gingivitis and periodontal diseases* | 3 | 2 |
| **H81** | *Disorders of vestibular function* | 1 | 0 |
| **J03** | *Acute tonsillitis* | 1 | 0 |
| **K62** | *Other diseases of anus and rectum* | 1 | 1 |
| **K85** | *Acute pancreatitis* | 1 | 5 |
| **M86** | *Osteomyelitis* | 1 | 0 |

**Legend.** (ICD-10-CM) International Classification of Diseases.

**TABLE 3S.** Characteristics of the acute episode and main outcomes for the entire population of HaH-HA and for the two study groups (n=441 each). Only the p-values ≤ 0.05 have been displayed.

|  | **Study Population (n=586)** | **HaH-HA (n=441)** | **p value** | **Controls (n=441)** | **p value** |
| --- | --- | --- | --- | --- | --- |
| **Total length of stay (days), mean (SD)** | 8.09 (4.88) | 7.89 (4.37) |  | 7.37 (6.17) |  |
| **Case Mix Index** | 0.68 | 0.69 |  | 0.73 |  |
| **Use of resources during Hospital Avoidance** |  | | |  | |
| All-cause Emergency Room visits, n (%) | 11 (1.87) | 6 (1.36) |  | N/A |  |
| All-cause In-Hospital re-admissions, n (%) | 28 (4.75) | 18 (4.08) |  | N/A |  |
| **Mortality during episode, n (%)** | 0 (0) | 0 (0) |  | 19 (4.31) | **N/A** |
| **Outcomes at 30 days after discharge** |  | | |  | |
| All-cause Emergency Room visits, n (%) | 36 (6.14) | 28 (6.35) |  | 34 (8.06) | **.032** |
| All-cause Hospital admissions |  | | |  | |
| Unplanned Hospital admissions, n (%) | 37 (6.31) | 24 (5.44) |  | 23 (5.45) |  |
| Planned admissions, n (%) | 19 (3.24) | 13 (2.95) |  | 10 (2.37) |  |
| Mortality, n(%) | 7 (1.19) | 7 (1.59) |  | 7 (1.66) |  |

**Legend.** CCA, Cost Consequence Analysis; Study population: all patients included in Hospital at Home-Hospital Avoidance (HaH-HA); HaH-HA, corresponds to the study population after propensity score matching (PSM); Controls, conventional hospitalizations, after PSM.

**Table 4S.** Operational Cost (in €) during the acute episode and expenses during 30-days after discharge for the entire population and the two study groups. Only the p-values ≤ 0.05 of the category totals have been displayed.

|  | **Study Population (n = 586)** | **HaH-HA (n = 441)** | **Controls (n = 441)** | **p value** |
| --- | --- | --- | --- | --- |
| **STAFF** | | | | |
| Department Head | 21,637 | 16,770 | 20,032 | **<**.001 |
| Nursing co-ordinator | 19,457 | 15,080 | 23,194 |  |
| Physicians | 127,022 | 98,415 | 104,983 |  |
| Resident Physicians | 0 | 0 | 101,166 |  |
| Registered nurses | 293,620 | 227,489 | 238,948 |  |
| Nursing assistants | 0 | 0 | 152,070 |  |
| Physiotherapists | 0 | 0 | 11,690 |  |
| Social workers | 0 | 0 | 11,690 |  |
| Secretary | 31,561 | 24,461 | 14,853 |  |
| **Staff** | **493,298** | **382,215** | **678,625** |  |
| **PHARMACOLOGICAL TREATMENT and NON-PHARMACOLOGICAL** | | | | |
| Anti-infective therapy | 59,694 | 36,296 | 27,488 |  |
| Other pharmacological treatment | 8,187 | 5,927 | 25,087 |  |
| Oxygen Therapy *(new prescription)* | 4,470 | 3,909 | N/A |  |
| Nebulizer Therapy *(new prescription)* | 2,515 | 2,314 | N/A |  |
| **Treatment** | **74,865** | **48,445** | **52,575** |  |
| **CONSUMABLES** | | | | |
| **Consumables** | **18,455** | **13,887** | **39,200** | <.001 |
| **DIAGNOSTIC TESTS** | | | | |
| Laboratory tests | 8,067 | 5,895 | 29,378 | <.001 |
| Diagnostic images | 4,706 | 3,190 | 25,194 |  |
| **Diagnostic Tests** | **12,773** | **9,085** | **54,572** |  |
| **STAFF TRANSPORTATION** | | | | |
| **Staff Transportation** | **21,286** | **16,017** | **N/A** | N/A |
| **CATERING** | | | | |
| **Catering** | **N/A** | **N/A** | **65,519** | N/A |
| **STRUCTURE** | | | | |
| **Structure** | **7,448** | **5,636** | **66,787** | <.001 |
| **TOTAL** | **628,126** | **475,286** | **957,279** | <.001 |
| **Transitional Care (30d after discharge)** | **515,094** | **337,360** | **451,078** | <.001 |

**Legend.** Study population: all patients included in Hospital at Home-Hospital Avoidance (HaH-HA); HaH-Ha, corresponds to the study population after propensity score matching (PSM); Controls, conventional hospitalizations, after PSM.

**Figure 1S – Results of the satisfaction questionnaire administered to HaH-HA patients/careers at discharge.**

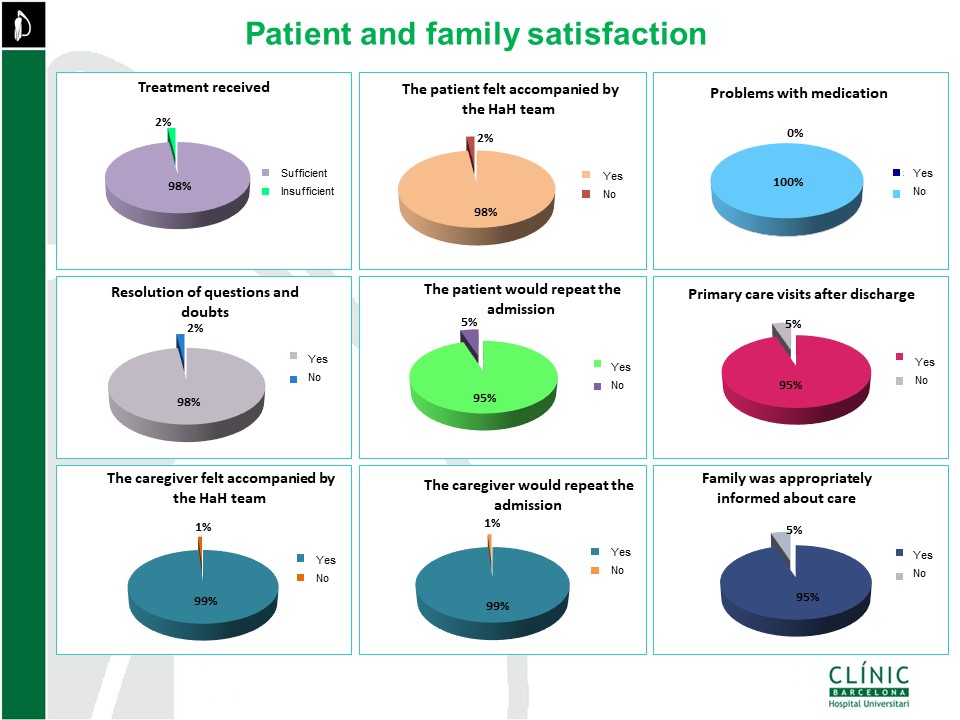
